# Antibodies against SARS-CoV-2 among health care workers in a country with low burden of COVID-19

**DOI:** 10.1101/2020.06.23.20137620

**Authors:** Mina Psichogiou, Andreas Karabinis, Ioanna D. Pavlopoulou, Dimitrios Basoulis, Konstantinos Petsios, Sotirios Roussos, Maria Patrikaki, Edison Jahaj, Konstantinos Protopapas, Konstantinos Leontis, Vasiliki Rapti, Anastasia Kotanidou, Anastasia Antoniadou, Garyphallia Poulakou, Dimitrios Paraskevis, Vana Sypsa, Angelos Hatzakis

## Abstract

Greece is a country with limited spread of SARS-CoV-2 and cumulative infection attack rate of 0.12% (95%CI 0.06%-0.26%). Health care workers (HCWs) are a well-recognized risk group for COVID-19. The study aimed to estimate the seroprevalence of antibodies to SARS-CoV-2 in two hospitals and assess potential risk factors. Hospital-1 was involved in the care of COVID-19 patients while hospital-2 was not. A validated, rapid, IgM/IgG antibody point-of care test was used. 1,495 individuals consented to participate (response rate 77%). The anti-SARS-CoV-2 weighted prevalence was 1.07% (95%CI 0.37-1.78) overall and 0.44% (95%CI 0.12-1.13) and 2.4% (95%CI 0.51-8.19) in hospital-1 and hospital-2, respectively. The overall, hospital-1, and hospital-2 seroprevalence was 9, 3 and 20 times higher than the estimated infection attack rate in general population, respectively. Suboptimal use of personal protective equipment was noted in both hospitals. These data have implications for the preparedness of a second wave of COVID-19 epidemic.

## Introduction

Coronavirus disease 2019 (COVID-2019) caused by a novel coronavirus [severe acute respiratory syndrome coronavirus-2 (SARS-CoV-2)] emerged in Wuhan, China in December 2019 (*1*) and spread worldwide in 212 countries and territories causing more than 5.8 million cases and 360,000 deaths within a period of 5 months (*1*).

In Greece, the first COVID-19 case was diagnosed on February 26. On March 23, a nation-wide lockdown was enforced to reduce ongoing virus transmission as a response to this pandemic. As of May 30, there were 2,915 confirmed cases and 175 related deaths in Greece with a death rate of 16 per 1,000,000 population, which is one of the lowest in Europe (*2*). Modelling data suggest that by the end of April 2020, when the first wave of epidemic was completed, the infection attack rate in Greece was 0.12% (95% Crl: 0.06-0.26%) which corresponds to 13,200 total infections (95% Crl: 6,206 – 27,700) and a case ascertainment rate of 19.1% (95 CI 9.1-40.6) (*3*).

Health care workers (HCWs) is a well-known risk group for coronavirus infections (*4,5*), accounted for a significant proportion of COVID-19 infections worldwide. By February 24, 2020, 3,387 HCWs out of 77,262 (4.4%) cases reported in China were HCWs (*6*). The majority of these HCWs were documented at Hubei province, the epicentre of the epidemic. In a comprehensive analysis of 9,684 HCWs from Tongji Hospital in Wuhan, Hubei, the symptomatic infection rate was 1.1% while the respective asymptomatic infection rate was estimated at 0.9%. Nurses held a higher infection risk than physicians [OR: 2.07 (95% CI 1.7-4.3)] (*7*). The city of Daegu, South Korea, had the first large outbreak of COVID-19 outside China. 121 HCWs were infected with infection rates 4.42 cases/1000 compared with 2.72 in the general population. Among HCWs, the infection rates were 2.37, 4.85, and 5.14 cases/1,000 among doctors, nurses, and nurse assistants, respectively (*8*). In a large study among HCWs from the Netherlands, of whom 9,705 were hospital employees, a total of 1,353 (14%) reported fever and respiratory symptoms. Of those, 86 (6%) were infected with SARS-CoV-2, representing 1% of all HCWs employed (*9*). Higher infection rates of SARS-CoV-2 by RT-PCR, ranging from 5-44%, were observed in HCWs from UK, Spain, Italy and US (*10-17*).

Serologic methods based on antibody testing (anti-SARS-CoV-2) could provide a more accurate estimate of epidemic size by detecting diagnosed and undiagnosed cases. Antibody methods rely on detection of IgM, IgG, IgA, or total antibodies by a variety of methods (*18,19*).

The prevalence of the SARS-CoV-2 antibodies among HCWs was assessed in a number of studies from countries with high burden of SARS-CoV-2 infection where the reported anti-SARS-CoV-2 seroprevalence ranges from 1.6 - 45.3% (*20-25*) Few studies used serological methods in the context of outbreak investigation (*26,27*).

This study aimed to assess the seroprevalence of antibodies to SARS-CoV-2 in HCWs of two Greek hospitals during the current epidemic and identify potential risk factors for infection.

## Patients and Methods

This cross-sectional study recruited HCWs aged more than 18 years from two hospitals. The designated hospital-1 is a 500-bed tertiary General Hospital providing care to COVID-19 patients. Hospital-2 with 134 beds is a Cardiac Surgery Center not involved in the care of COVID-19. The eligible personnel was in total 1,952, 1,120 in hospital-1, and 832 in hospital-2.

Two groups were investigated 1) first-line health care workers (FL-HCWs), defined as personnel whose activities involve contact with patients, and 2) second-line health care workers (SL-HCWs), such as office employees, technical personnel, cleaning personnel etc.

Testing was offered at one specified location in each hospital for a period of 4 weeks, 13 April-14 May 2020, and 30 April – 15 May 2020 in hospital-1 and −2, respectively. Informed consent was obtained from all participants who were interviewed using a structured questionnaire including demographics, education, position within hospital, exposure to COVID-19, use of personal protective equipment (PPE), and symptoms related to COVID-19. The data were directly recorded in a secure database. All participants were immediately informed on their test results, and they were offered a short posttest counseling session. The study was approved by the Institutional Review Board of both hospitals.

Testing was based on the GeneBody COVID-19 IgM/IgG detection, which is a chromatographic immunoassay for the rapid and differential test of immunoglobulin M and immunoglobulin G against SARS-CoV-2. Serologic testing for SARS-CoV-2 antibodies was performed using capillary blood according to the manufacturer’s instructions (Genebody Inc.). Samples were concluded as reactive if the IgM or the IgG or both bands were positive using a colorimetric reader (Confiscope G20 analyser). Positive individuals were immediately retested and the concordant were considered positive.

The antibody assay was validated in a serological panel of 107 hospitalized, symptomatic, positive by RT-PCR, COVID-19 patients (Panel A) and in a second panel (Panel B) including 150 samples collected before SARS-CoV-2 epidemic (see Appendix).

To calculate the prevalence of antibodies to SARS-CoV-2, firstly, we calculated the unweighted proportions of positive testsand then we obtained the prevalence after weighting for the age distribution of the adult population (18-69 years old) in Athens Metropolitan area from the 2011 census. Secondlywe adjusted the weighted proportion for the sensitivity and specificity of the test, as assessed from the validation in the serological panels A and B, using the epiR package (R version 3.6.3, R Foundation for Statistical Computing, Vienna, Austria).

## Results

A total of 1,495 HCWs consented to participate. The overall participation rate was 77% (81% and 71% in hospitals-1 and 2, respectively). Of 1,495 individuals tested, 69.7% were women, 61.7% were aged 35 to 54 years old, with a mean age (SD) of 46.4(10.3) years. FL-HCWs accounted for 73.4 % of those tested. Subjects’ characteristics are listed in **Table**.

**Table 1:** Socio-demographic characteristics and weighted prevalence of anti-SARS CoV-2 of 1,495 participants in two hospitals in Athens.

| **Covariate** | **Population**<br>**(N)** | **Anti-SARS-**<br>**CoV-2 (+)** | **Weighted**<br>**prevalence**<br>**with 95% CI<sup>1</sup>** |
| --- | --- | --- | --- |
| Overall | 1,495 | 15 | 1.07 (0.37, 2.78) |
| Hospital |  |  |  |
| Hospital 1 | 906 | 4 | 0.44 (0.12, 1.13) |
| Hospital 2 | 589 | 11 | 2.40 (0.51, 8.19) |
| Gender |  |  |  |
| Male | 453 | 5 | 1.36 (0.11, 5.90) |
| Female | 1,042 | 10 | 0.97 (0.31, 3.14) |
| Age (y) |  |  |  |
| 18-34 | 231 | 1 | 0.50 (0.01, 2.75) |
| 35-54 | 922 | 8 | 1.00 (0.43, 1.96) |
| 55-70 | 342 | 6 | 2.02 (0.74, 4.34) |
| Country of birth |  |  |  |
| Greece | 1,355 | 14 | 1.10 (0.36, 2.94) |
| Other | 140 | 1 | 0.67 (0.02, 14.50) |
| Marital status |  |  |  |
| Married | 910 | 11 | 1.01 (0.35, 5.99) |
| Divorced / widowed | 134 | 0 | 0.00 (0.00, 4.04) |
| Single | 451 | 4 | 0.89 (0.08, 6.52) |
| Members of household |  |  |  |
| 1 | 260 | 2 | 0.91 (0.02, 6.79) |
| 2 | 407 | 2 | 0.65 (0.02, 4.40) |
| 3 | 313 | 5 | 1.53 (0.28, 9.24) |
| 4 | 383 | 5 | 1.32 (0.20, 10.46) |
| 5+ | 132 | 1 | 0.55 (0.01, 16.92) |
| Highest completed level of education |  |  |  |
| Master's degree/Doctorate | 416 | 2 | 0.39 (0.05, 4.93) |
| University or equivalent | 632 | 10 | 1.81 (0.47, 5.18) |
| Technical education or below | 447 | 3 | 0.56 (0.05, 8.25) |
| Job title |  |  |  |
| Healthcare workers | 1,097 | 11 | 1.00 (0.50, 1.79) |
| Nonhealthcare workers | 398 | 4 | 1.01 (0.27, 2.55) |
| Symptoms<sup>2</sup> |  |  |  |
| Any symptom | 150 | 3 | 2.02 (0.15, 13.78) |
| No symptom | 1,345 | 12 | 0.98 (0.28, 2.80) |

<sup>1</sup> *Weighted prevalence for age and test performance*
<sup>2</sup> *Among fever, cough, and shortness of breath*

A total of 15 individuals tested positive for anti-SARS-CoV-2, eleven of them for IgG only, three for IgM only and one for both IgM/IgG. After adjusting for age and test performance-assuming 87% sensitivity and 100% specificity the weighted seroprevalence for anti-SARS-CoV-2 in the total population was 1.07% (95%CI 0.37, 2.78). The weighed seroprevalence in hospital-1 was 0.44% (95%CI 0.12, 1.13) and in hospital-2 2.40% (95%CI 0.51, 8.19) **(Table)**. The seroprevalence was 9, 3 times and 20 times higher in the overall hospital population, in hospital-1 and in hospital-2, respectively compared with the general population (0.12%, Crl:0.06-0.26) (*3*). No significant associations were noted in the seroprevalence according to gender, country of birth, education, number of members in the household, FL-HCWs, SL-HCWs and use of PPE. Anti-SARS-CoV-2 prevalence was higher with increasing age, but the trend was not statistically significant (p=0.10). The use of PPE was suboptimal in both hospitals. In hospital-1 and among the personnel treating COVID-19 the use of gloves, masks, glasses, gown was 96%, 99%, 56% and 63%, respectively. In hospital-2 the use of gloves and mask was reported in 99.7% and 100% while the use of glasses and gown occasionally (15%).

Among all participants, 150 (10.1%) reported some symptoms indicative of COVID-19 in the previous 3 months; 82 reported fever, and 111 of them cough; 27 reported shortness of breath. Overall, 1,345 (89.9%) reported no symptoms. The prevalence of anti-SARS-CoV-2 was 2.02% (95%CI 0.15, 13.78) and 0.98% (95%CI 0.28, 2.80) in those who reported and those not reporting symptoms, respectively but the difference was not statistically significant.

## Discussion

In this survey of SARS-CoV-2 antibodies among hospital personnel, the overall seroprevalence was 1.07% (95% CI 0.37, 2.78) using a validated point-of care assay. This low seroprevalence rate is consistent with the low burden of COVID-19 in Greece. However, in the total hospital population and in that of hospital-2, it was 9 and 20 times higher, respectively, compared to the cumulative infection attack rate estimated by mathematical modeling for the general population in Greece (*3*). This is not surprising since the spread of SARS-CoV-2 is highly heterogeneous (*28*). In New York State the prevalence of anti-SARS-CoV-2 was found 14.0% with a range of 3.6-22.7% (*28*).

Due to the low burden of infection, the study is underpowered for pointing out risk factors. The difference in the prevalence between hospital-1 [0.44% (95%CI 0.12, 1.13)] and hospital-2 [2.40% (95%CI 0.51, 8.19)] is not significant. However, it is consistent with data suggesting that HCWs in hospitals involved in COVID-19 care could have a lower burden of infection than those not participating in COVID-19 care (*7, 21*). This is probably due to the use of PPE, which is the main determinant for risk of SARS-CoV-2 infection in the health care environment (*30*). In this study the use of PPE was suboptimal in both hospitals. Other reported risk factors are working in high-risk departments, long duty hours, practicing suboptimal hand hygiene (*31*). Of the 42,600 HCWs caring for COVID-19 patients in the second half of the China epidemic, none was infected, suggesting that sufficient precautions and rigorous enforcement of PPE are the major determinants for eliminating COVID-19 infection (*6*).

A further challenge is whether SARS-CoV-2 infection can be truly attributed to hospital-acquired infections, especially in countries with a high burden of community infection (*28*). In the study of Lai Y et al, contact with patients (59%), colleagues with infection (11%), and community acquired infection (13%) were the main routes of exposure among HCWs (*7*).

Contradicting results are noted in two large studies from Madrid and Birmingham. The anti-SARS-CoV-2 prevalence is higher in HCWs working in areas with exposure to COVID-19 (31-34%) compared with low-risk area (26%) and external workers (30%) in Madrid (*24*). On the contrary in Birmingham study the anti-SARS-CoV-2 prevalence was higher among general medicine and housekeeping general personnel (30-35%) compared with intensive care and emergency medicine (13-15%) (*21*).

Several study limitations are noted: 1) The sensitivity of the currently existing antibody assays is not well known since they were registered using convalescent sera from symptomatic hospitalized patients and their sensitivity was not assessed in asymptomatic or mildly symptomatic patients (*18,19*). 2) At present, data on post-infection immunity are lacking. Studies from the previous SARS-CoV-1 outbreak have shown a steady prevalence decrease with time (*18,19*) 3) Higher antibody titers are associated with infection severity (*18,19*). 4) The study, due to the low anti-SARS-CoV-2 prevalence, is underpowered to detect risk factors. 5) The prevalence of anti-SARS-CoV-2 in the Greek population is not known and the infection attack rate estimated from a modelling study was used as surrogate of the general population prevalence. 6) Several studies, non-peer reviewed available as preprint, were used.

In conclusion, the burden of SARS-CoV-2 infection among hospital personnel in Athens is low, consistent with the low burden of infection in the country. The use of PPE was suboptimal. These findings have implications for the preparedness of a second wave of COVID-19.

## Data Availability

Data will be available at the Pergamos database of the National and Kapodistrian University of Athens, upon acceptance of the article by a journal

## The authors would like to thank

Panayiotis Axaopoulos, Georgios Goumas, Evangelos Kokolesis, Michaella Alexandrou, Sofia Radi, Dimitra Siakali, Erica Alexandrou, Dimosthenis Theodosiadis, Ilias Sinanidis, Charalambos Kazamiakis, as well as Onassis C.S.C. staff members: George Stravopodis & Sofia Hatzianastasiou, and the Advisory board members in Laiko General Hospital: John Boletis, Nikolaos Sypsas, Michalis Samarkos, Ioannis Floros, Theoni Zougkou, Amalia Karapanou, Michalis Sambanis

